# Risk Factors for Progressive Cardiorenal Syndrome Events in Patients with Chronic Kidney Disease: The CRIC Study

**DOI:** 10.64898/2026.10.30.26366964

**Authors:** Meredith C McAdams, Jiang He, Bernard Jaar, Hua He, Cong Zhao, Siyi Geng, Amanda H. Anderson, Vecihi Batuman, Joshua D. Bundy, Jon Taliercio, Panduranga Rao, Sarah Schrauben, Milica Djokic, Joel Merriman, Giselle Sosa, Alan Go, James P Lash, L Lee Hamm, Jing Chen, the CRIC Study Investigators

## Abstract

**Background:** Cardiorenal syndrome (CRS) is associated with poor outcomes. Risk factors for progressive CRS are not understood in patients with chronic kidney disease (CKD).

**Objectives:** This study aimed to determine the incidence of progressive CRS and risk factors in individuals with CKD.

**Methods:** Adults with CKD in the Chronic Renal Insufficiency Cohort (CRIC) study excluding those with heart failure (HF) at baseline were included (n = 3,557). We defined progressive CRS as developing HF and end-stage kidney disease (ESKD) within 12 months. ESKD was defined as needing renal replacement therapy or renal transplant. The associations of baseline risk factors with progressive CRS were examined using Cox proportional hazards models. A sensitivity analysis assessed risk factors in those who developed HF before ESKD and vice versa.

**Results:** 161 patients (4.5%) developed progressive CRS. Traditional risk factors [multivariable-adjusted hazard ratios (HR) (95% CI)] associated with progressive CRS in multivariable models included lower education [1.90 (1.28,2.83)], cardiovascular disease [1.53 (1.07,2.18)], lower estimated glomerular filtration rate [0.64 (0.51,0.82)] and higher albumin to creatinine ratio [2.44 (1.98,3.02)]. Novel risk factors associated included higher levels of alkaline phosphatase, brain natriuretic peptide (BNP), tumor necrosis factor alpha (TNF-α), growth differentiation factor 15 (GDF-15), hemoglobin A1C, and lower hemoglobin. Risk factors associated with both ESKD before HF and HF before ESKD were kidney dysfunction, albuminuria, and mineral bone disorder (higher alkaline phosphatase and PTH), inflammation (TNF-α), fibrotic factor (GDF-15), and anemia. Diabetes, CVD, systolic blood pressure, waist circumference, HbA1c, and BNP were more strongly associated when HF preceded ESKD.

**Conclusions:** Both traditional risk factors and novel risk factors associated with inflammation, mineral bone disorder, anemia, diabetes control, albuminuria, worse kidney function, and fibrotic factors are associated with progressive CRS in individuals with CKD. Many of these risk factors are modifiable representing potential areas for intervention.

## Introduction

The overlapping and nuanced relationship between the heart and kidneys is encompassed by the term cardiorenal syndrome (CRS). The overlap of these systems involves various physiologic entities including hemodynamics, neurohormonal activation and inflammation. CRS is common with anywhere from 30-50% of patients with HF having moderate to severe kidney dysfunction (1–5). Chronic kidney disease (CKD) is an independent risk factor for cardiovascular disease (CVD), and cardiovascular disease is widely recognized as the most common cause of mortality in individuals with both CKD and end-stage kidney disease (ESKD) (6,7). Concurrent cardiac and renal disease is associated with worse outcomes. One meta-analysis showed that in patients with HF having any renal impairment increased all-cause mortality compared to having no renal impairment. This study also showed that mortality risk increased as renal function decreased, with a 15% increased risk for every 0.5 mg/dl increase in creatinine and a 7% increased risk for every 10 ml/min decrease in estimated glomerular filtration rate (eGFR) (5). Individuals hospitalized with heart failure who had either CKD, or an AKI are at increased risk for in-hospital mortality (8). It has also been shown that worsening renal function during or after discharge in patients hospitalized with HF is associated with increased mortality and increased risk of hospital readmission (9).

In patients with underlying CKD who experience acute HF, outcomes are also worse when compared to those with no underlying CKD. There have been some studies showing that individuals with CKD experienced higher rates of HF hospitalizations and higher rates of 30-day readmissions after HF hospitalization (10). Further it has been shown that patients with more advanced stages of CKD have higher rates of complications including AKI requiring dialysis, gastrointestinal bleeding and respiratory failure when hospitalized with HF (11). While it has been established that individuals with CKD and HF are at increased risk for mortality and hospitalizations, less is known regarding shared risk factors for progressive kidney function decline and heart failure in the CKD population. We aimed to evaluate the incidence and risk factors for progressive CRS in a multi-center diverse CKD population.

## Methods

### Population

The CRIC study included 3,939 patients between the ages of 21 to 74 with mild-to-moderate CKD (eGFR entry criteria was 20-30 mL/min/1.73m^2^) between May 2003 and August 2008 from seven clinical centers in the US (Ann Arbor/Detroit, MI; Baltimore, MD; Chicago, IL; Cleveland, OH; New Orleans, LA; Philadelphia, PA; and Oakland, CA) (12). Full inclusion and exclusion criteria have been previously described (13). Patients with ESKD on dialysis and those with kidney transplants were excluded. CRIC also excluded any participant who had advanced HF (New Your Heart Association Class III or IV) at baseline. All CRIC participants provided written consent for the study. The study protocol was approved by each institutional review board at each site. The CRIC study conformed to the Health Insurance Portability and Accountability Act (HIPPAA) guidelines. 3,557 participants were included in this analysis after excluding those with heart failure at baseline visits.

### Data Collection

Data was collected by trained CRIC staff during baseline and annual visits. The data collections were conducted using standardized methods (12). Questionnaires were used to collect baseline demographics, medical history, medications, and lifestyle risk factors. Standard methods were used to measure height, weight, waist circumference, and blood pressure. The CKD-EPI 2021 non-race-based equation was used to calculate estimated glomerular filtration rate (eGFR)(14). A 24-hour urine collection was collected and measured by radioimmunoassay. All laboratory measurements were conducted at the University of Pennsylvania CRIC Study central laboratory with stringent quality control.

### Exposure variables

We analyzed various available variables for association with the primary outcome. Traditional cardiovascular disease (CVD) risk factors evaluated included physical activity, smoking, alcohol use, history of clinical CVD, body mass index (BMI), waist circumference, systolic blood pressure, use of antihypertensive medications, history of diabetes mellitus, cholesterol levels, use to statin medications, eGFR, and albuminuria. We also evaluated novel risk factors including: calcium, phosphate, albumin, parathyroid hormone (PTH), fibroblast growth factor 23 (FGF23), myeloperoxidase, uric acid, homocysteine, 24-hour urine sodium, 24-hour urine potassium, aldosterone, high sensitivity C-reactive protein (hs-CRP), B-type natriuretic peptide (BNP), ST-2, galactin 3, connective tissue growth factor (CTGF), homeostatic model assessment (HOMA) insulin resistance, urine neutrophil gelatinase-associated Lipocalin-1 (NGAL-1), bioelectrical impedance analysis (BIA) fat free mass, fetuin A, interleukin-6 (IL-6), tumor necrosis factor alpha (TNF-α), interleukin 1 beta (IL-1β), fibrinogen, Fractalkine, chemokine ligand 12 (CXCL 12), growth differentiation factor 15 (GDF-15), HbA1C, and hemoglobin.

### Outcomes

Participants were contacted biannually and updated information on medical history and hospitalizations was obtained. Our outcome of interest was progressive CRS which we defined as development of both HF and end-stage kidney disease (ESKD), requiring dialysis or kidney transplant, within twelve months of each other. Progressive CRS type 2 was defined as having HF preceded ESKD, and progressive CRS type 4 was defined as having ESKD preceded HF. HF was identified using queries of hospitalization records; discharge codes were identified and codes relevant to heart failure led to retrieval of medical records for adjudicated review. The potential heart failure event records were reviewed by at least two study physicians and were confirmed when both reviewers agreed on the diagnosis. ESKD was determined by self-report from participants, review of medical records, and linkage to the US Renal Data System(15).

### Statistical Analysis

Baseline characteristics of the cohort were summarized by means with standard deviations for continuous variables, median with inter-quartile ranges for skewed variables or percentages for categorical variables, these were presented based on presence or absence of progressive CRS. Two-sample t-tests or Wicoxon rank-sum tests were used to test statistical significant as appropriate. χ2 was used to test significance for categorial variables. In the event of skewed variables logarithmic transformations were performed to normalize distributions and stabilize variances.

Cox proportional hazard models were used to assess the association of both traditional and novel risk factors with progressive CRS. The traditional risk factor models were adjusted for age, sex, race, and clinic site. Backward elimination was then used to select final models and used only those risk factors that were significant were retained. The novel risk factor models were adjusted for age, sex, race, and clinic site, high school education, current smoking CVD, diabetes, waist circumference, systolic BP, total cholesterol, low density lipoprotein (LDL), triglycerides, eGFR, and ACR. In sensitivity analyses, risk factors for CRS type 2 and CRS type 4 were evaluated. Kaplan-Meier curves were generated to compare survival among those who experienced the outcome of interest and those who did not.

## Results

A total of 161 patients of the cohort, or 4.5%, developed progressive cardiorenal syndrome. The baseline characteristics of those who developed cardiorenal syndrome and those who did not are shown in **Table 1**. The group who developed progressive CRS had an average age of 58.5 years and those without CRS had an average age of 57.3 years. Those who developed progressive CRS were more likely to be black or Hispanic than those without. Those who developed progressive CRS were more likely to have history of clinical CVD, diabetes, hypertension. Those individuals who developed progressive CRS also had a statistically significantly higher systolic blood pressure, hemoglobin A1C, serum phosphate, alkaline phosphatase, parathyroid hormone, FGF-23, urine albumin to creatinine ratio, and BNP level compared to those who did not develop CRS. The progressive CRS group had a statistically significantly lower serum albumin and hemoglobin level compared to the non-CRS group. Rates of weekly physical activity, history of atrial fibrillation, use of antihypertensive medications, and use of statins were not different between the groups. The age-adjusted incidence of progressive CRS by sex and race and ethnicity is shown in **Figure 1**. The overall incidence of progressive CRS was 4.00 per 1000 person-year for women and 4.96 for men. Hispanics had the highest incidence with an incidence of 6.40 for women and 7.69 for men, the incidence was lowest in whites with an incidence of 1.83 for women and 3.50 for men. Men had a higher incidence in all races and ethnicities except for the “other races” category.

**Figure 1.**
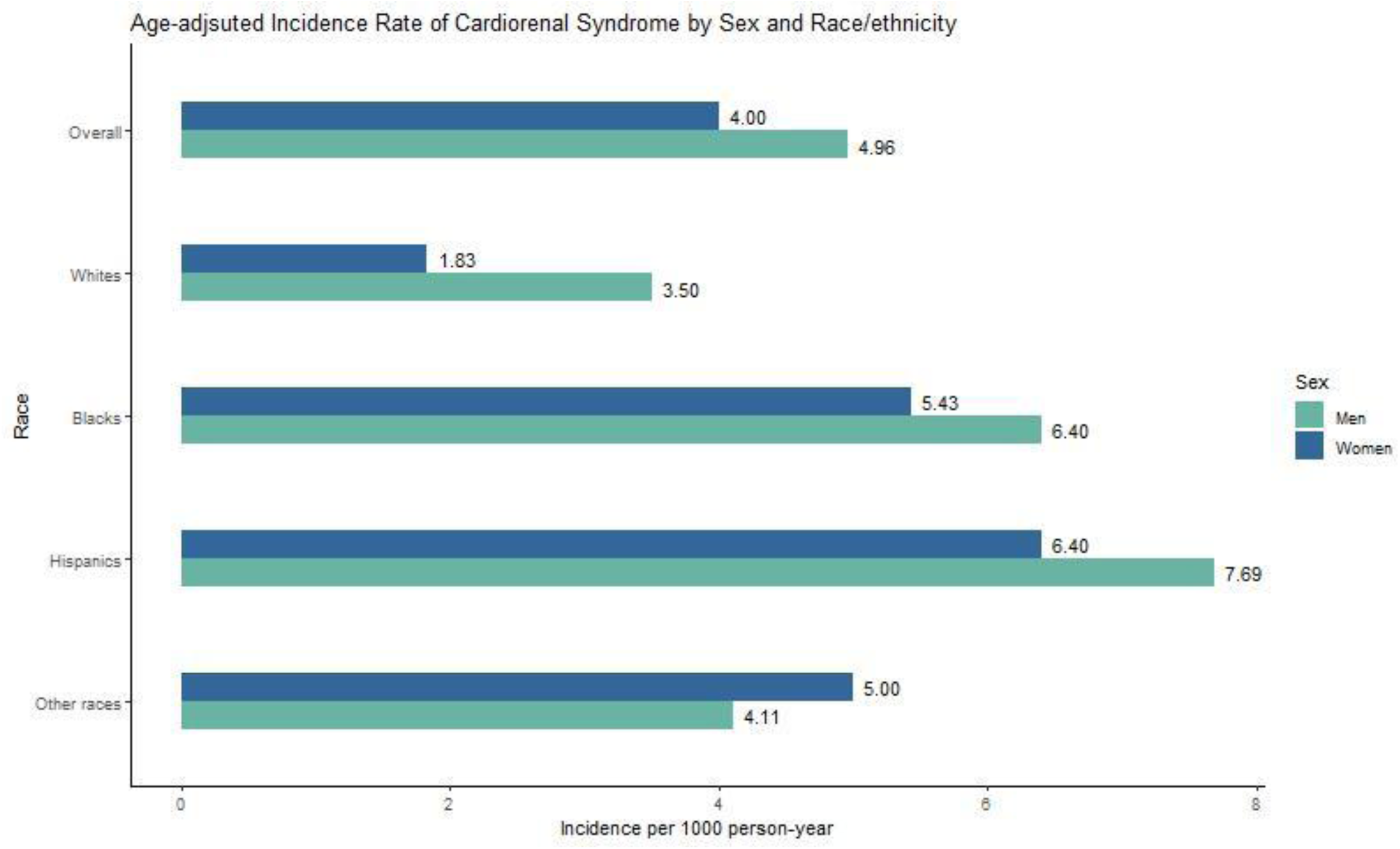
Age-adjusted incidence of progressive cardiorenal syndrome by sex and race/ethnicity. Incidence is reported per 1000 person-year.

**Table 1.** Baseline characteristics of study participants with and without progressive cardiorenal syndrome.

|  | **Progressive cardiorenal syndrome (N=161)** | **No cardiorenal syndrome (N= 3396)** | ***P* value** |
|---|---|---|---|
| Age, mean (SD), years | 58.5 (10.2) | 57.3 (11.2) | 0.19 |
| Male, no. (%) | 94 (58.4) | 1847 (54.4) | 0.36 |
| Race/ethnicity, no. (%) | - | - | - |
| Non-Hispanic white | 45 (28.0) | 1476 (43.5) | 0.001 |
| Non-Hispanic black | 81 (50.3) | 1352 (39.8) |  |
| Hispanic | 29 (18.0) | 431 (12.7) |  |
| Other | 6 (3.7) | 137 (4.0) |  |
| High school graduate, no. (%) | 104 (64.6) | 2720 (80.1) | <0.001 |
| Physical activity, mean (SD), MET/week | 187.7 (142.7) | 203.0 (147.7) | 0.20 |
| Current smoking, no. (%) | 30 (18.6) | 433 (12.8) | 0.04 |
| Weekly alcohol drinking, no. (%) | 89 (55.3) | 2189 (64.5) | 0.02 |
| Body mass index, mean (SD), kg/m<sup>2</sup> | 32.5 (9.1) | 31.9 (7.6) | 0.27 |
| Waist circumference, mean (SD), cm | 106.7 (20.6) | 105.3 (17.3) | 0.32 |
| History of clinical CVD, no. (%) | 62 (38.5) | 872 (25.7) | <0.001 |
| History of hypertension, no. (%) | 153 (95.0) | 2891 (85.1) | <0.001 |
| History of diabetes, no. (%) | 104 (64.6) | 1541 (45.4) | <0.001 |
| History of atrial fibrillation, no. (%) | 25 (15.5) | 457 (13.5) | 0.53 |
| Use of antihypertensive medications, no. (%) | 151 (98.1) | 2829 (96.8) | 0.51 |
| Use of statins, no. (%) | 95 (59.7) | 1778 (52.7) | 0.099 |
| Systolic BP, mean (SD), mm Hg | 137.3 (22.4) | 128.0 (21.9) | <0.001 |
| Hemoglobin A1C, mean (SD), mg/dL | 7.3 (2.0) | 6.6 (1.5) | <0.001 |
| HDL cholesterol, mean (SD), mg/dL | 45.4 (12.9) | 48.0 (15.7) | 0.04 |
| LDL cholesterol, mean (SD), mg/dL | 106.8 (38.1) | 103.7 (35.3) | 0.28 |
| Triglycerides, median (inter-quartile range), mg/dL | 140.0 [96.0, 186.0] | 127.0 [88.0, 183.0] | 0.06 |
| eGFR, mean (SD), ml/min/1.73m<sup>2</sup> | 35.8 (13.4) | 45.3 (15.3) | <0.001 |
| ACR, median (inter-quartile range), ug/mg | 601.1 [127.6, 1829.1] | 42.9 [7.9, 392.8] | <0.001 |

SD, standard deviation. No, number. MET, metabolic equivalent. Kg, kilogram. M<sup>2</sup>, meters squared. Cm, centimeters. CVD, cardiovascular disease. BP, blood pressure. A1C, glycated hemoglobin. HDL, high-density lipoprotein. LDL, low-density lipoprotein. eGFR, estimated glomerular filtration rate. ACR, albumin to creatinine ratio.

Traditional risk factors associated with progressive CRS are presented in **Table 2**. Traditional risk factors significantly associated with development of progressive CRS included not having graduated from high school, current smoking, history of clinical CVD, higher systolic blood pressure, history of diabetes, higher total cholesterol, higher log triglycerides, lower eGFR, and higher log ACR. Risk factors that remained statistically significant in the multivariable backward selection model included less than high school education, history of clinical CVD, lower eGFR, and higher log ACR.

**Table 2.** Hazard ratios of progressive cardiorenal syndrome associated with traditional risk factors.

|  | **Age, sex, race, and clinic site-adjusted** |  | **Multivariable-adjusted\*** |  |
|---|---|---|---|---|
|  | **Hazard ratio (95%CI)** | ***P*-value** | **Hazard ratio (95%CI)** | ***P*-value** |
| Not graduated from high school | 1.81 (1.24, 2.64) | 0.0020 | 1.90 (1.28, 2.83) | 0.002 |
| Log (Physical activity) SD:0.73 MET/week | 0.90 (0.78, 1.05) | 0.1906 | - | - |
| Current smoking | 1.57 (1.04, 2.37) | 0.0305 | - | - |
| Weekly alcohol drinking | 0.78 (0.56, 1.08) | 0.1298 | - | - |
| History of clinical CVD | 1.85 (1.33, 2.58) | 0.0003 | 1.53 (1.07, 2.18) | 0.02 |
| Body mass index SD:7.68 kg/m<sup>2</sup> | 1.06 (0.90, 1.25) | 0.4721 | - | - |
| Waist circumference SD: 17.41 cm | 1.08 (0.92, 1.28) | 0.3313 | - | - |
| Systolic blood pressure SD: 22.01 mmHg | 1.45 (1.25, 1.68) | <.0001 | - | - |
| Use of antihypertensive medication | 1.54 (0.49, 4.87) | 0.4598 | - | - |
| History of diabetes | 2.15 (1.54, 3.01) | <.0001 | - | - |
| Total cholesterol SD:45.5 mg/dL | 1.18 (1.02, 1.38) | 0.0255 | - | - |
| HDL cholesterol SD:15.59 mg/dL | 0.86 (0.71, 1.04) | 0.1208 | - | - |
| LDL cholesterol SD:35.46 mg/dL | 1.12 (0.96, 1.31) | 0.1395 | - | - |
| Log (Triglycerides) SD:0.55 mg/dL | 1.18 (1.01, 1.38) | 0.037 | - | - |
| Use of statins | 1.28 (0.93, 1.78) | 0.1327 | - | - |
| eGFR SD:15.30 ml/min/1.73m<sup>2</sup> | 0.45 (0.37, 0.56) | <.0001 | 0.64 (0.51, 0.82) | <0.001 |
| Log (ACR) SD:2.3 ug/mg | 2.97 (2.45, 3.61) | <.0001 | 2.44 (1.98, 3.02) | <0.001 |

\* Backward elimination method was used to select final model and only traditional risk factors that were significant (p<0.05) were retained. Age, sex, race, and clinic sites included.
MET, metabolic equivalent. CVD, cardiovascular disease. SD, standard deviation. HDL, high-density lipoprotein. LDL, low-density lipoprotein. eGFR, estimated glomerular filtration rate. ACR, albumin to creatinine ratio.

The association of novel risk factors with progressive CRS was presented in **Table 3**. Multiple novel risk factors were statistically associated with progressive CRS in the age, sex, race, and clinic adjusted model, including lower albumin, higher phosphate, lower hemoglobin, higher PTH, and higher values of FGF23, BNP, ST-2, CTGF, urine NGAL-1, IL-6, TNF-α, and GDF-15. In multivariable-adjusted models, each 1-SD increase in BNP, TNF-α, GDF-15, hemoglobin A1c, and alkaline phosphatase was associated with approximately 20%–35% higher risk of progressive CRS, whereas each 1-SD increase in hemoglobin was associated with a 20% lower risk.

**Table 3.** Multivariable-Adjusted Hazard Ratios for Progressive Cardiorenal Syndrome per 1-SD Increase in Novel Risk Factors.

|  | **Age, sex, race, and clinic site-adjusted** |  | **Multivariable-adjusted<sup>T</sup>** |  |
|---|---|---|---|---|
|  | **Hazard ratio (95%CI)** | ***P*-value** | **Hazard ratio (95%CI)** | ***P*-value** |
| serum albumin SD:0.47<br>g/dL | 0.59 (0.51, 0.68) | <.0001 | 0.90 (0.75, 1.07) | 0.23 |
| serum phosphate SD:0.66<br>mg/dL | 1.46 (1.27, 1.68) | <.0001 | 1.09 (0.92, 1.29) | 0.33 |
| serum calcium SD:0.46<br>mg/dL | 1.02 (0.86, 1.20) | 0.8585 | 0.95 (0.80, 1.12) | 0.54 |
| **Log (alkaline phosphatase) SD:0.32**<br>**U/L** | 1.42 (1.23, 1.64) | <.0001 | 1.19 (1.01, 1.41) | 0.04 |
| Log (PTH) SD: 0.70<br>pg/mL | 1.71 (1.46, 2.01) | <.0001 | 1.22 (1.00, 1.48) | 0.05 |
| Log (FGF23) SD:0.74<br>RU/mL | 1.61 (1.39, 1.86) | <.0001 | 1.10 (0.90, 1.34) | 0.35 |
| Log (Myeloperoxidase) SD:0.82<br>pmol/L | 1.13 (0.95, 1.34) | 0.1706 | 1.01 (0.86, 1.19) | 0.90 |
| uric acid SD:1.88<br>mg/dL | 1.01 (0.86, 1.19) | 0.8733 | 0.88 (0.73, 1.07) | 0.19 |
| Log (Homocysteine) SD:0.35<br>umol/L | 1.26 (1.06, 1.49) | 0.0087 | 0.99 (0.80, 1.22) | 0.92 |
| 24-hour urine sodium SD:77.15<br>mmol/24 hours | 1.04 (0.88, 1.23) | 0.6304 | 1.08 (0.91, 1.28) | 0.40 |
| 24-hour urine potassium SD:26.27 mmol/24 hours | 0.82 (0.67, 1.01) | 0.0573 | 0.86 (0.69, 1.07) | 0.18 |
| Log (Aldosterone) SD:0.69<br>pg/mL | 1.06 (0.90, 1.24) | 0.4885 | 1.02 (0.85, 1.22) | 0.84 |
| **Log (BNP) SD:1.25**<br>pg/mL | 1.82 (1.55, 2.13) | <.0001 | 1.35 (1.12, 1.63) | 0.002 |
| Log (ST-2) SD:0.56<br>ng/mL | 1.32 (1.11, 1.57) | 0.0018 | 1.06 (0.89, 1.26) | 0.50 |
| galectin 3 SD:7.34<br>ng/mL | 1.34 (1.20, 1.50) | <.0001 | 1.11 (0.97, 1.27) | 0.13 |
| Log (CTGF) SD:1.15<br>ng/mL | 1.94 (1.60, 2.36) | <.0001 | 1.19 (0.96, 1.47) | 0.12 |
| Log (HOMA-insulin resistance) SD:0.83<br>mmol/L * uU/mL | 1.21 (1.03, 1.41) | 0.0199 | 1.03 (0.85, 1.25) | 0.76 |
| Log (urine NGAL-1) SD:1.24<br>ng/mL | 1.73 (1.50, 2.01) | <.0001 | 1.05 (0.87, 1.28) | 0.60 |
| Log (BIA fat free mass) SD:0.27<br>° | 0.80 (0.69, 0.94) | 0.0063 | 0.93 (0.79, 1.09) | 0.37 |
| fetuin A SD:111493.74<br>ng/mL | 0.95 (0.81, 1.12) | 0.5591 | 1.02 (0.86, 1.21) | 0.84 |
| Log (IL-6) SD:0.89<br>pg/mL | 1.28 (1.11, 1.46) | 0.0005 | 1.12 (0.94, 1.32) | 0.20 |
| **Log (TNF-α) SD:0.72**<br>pg/mL | 1.41 (1.25, 1.60) | <.0001 | 1.24 (1.04, 1.47) | 0.01 |
| Log (IL-1β) SD:1.70<br>pg/mL | 1.05 (0.89, 1.24) | 0.5621 | 1.04 (0.87, 1.23) | 0.70 |
| Log (TGF-β) SD:0.78<br>ng/ML | 0.97 (0.82, 1.15) | 0.7292 | 1.01 (0.85, 1.21) | 0.87 |
| Fibrinogen SD:1.19<br>g/L | 1.54 (1.33, 1.77) | <.0001 | 1.05 (0.90, 1.24) | 0.52 |
| Fractalkine SD:0.37<br>pg/ml | 1.36 (1.20, 1.55) | <.0001 | 1.04 (0.89, 1.23) | 0.61 |
| CXCL 12 SD:550.26<br>pg/ml | 1.42 (1.23, 1.63) | <.0001 | 1.12 (0.95, 1.31) | 0.19 |
| Log (hsCRP) SD:1.26<br>mg/L | 1.06 (0.91, 1.25) | 0.4471 | 1.11 (0.93, 1.32) | 0.24 |
| **Log (GDF-15) SD:0.60**<br>pg/mL | 2.17 (1.81, 2.59) | <.0001 | 1.35 (1.05, 1.73) | 0.02 |
| **HbA1c SD: 1.53**<br>**%** | 1.49 (1.32, 1.69) | <.0001 | 1.35 (1.12, 1.62) | 0.002 |
| **CBC hemoglobin SD:1.77 g/dL** | 0.64 (0.54, 0.77) | <.0001 | 0.80 (0.65, 0.97) | 0.03 |

<sup>T</sup> Adjusted for age, sex, race, clinic sites, high school education, current smoking, CVD, diabetes, waist circumference, systolic BP, total cholesterol, LDL, triglycerides, eGFR, and ACR.
SD, standard deviation. PTH, parathyroid hormone. FGF23, fibroblast growth factor 23. BNP, B-type natriuretic peptide. CTGF, connective tissue growth factor. HOMA, homeostatic model assessment. NGAL-1, neutrophil gelatinase-associated Lipocalin-1. BIA, bioelectrical impedance analysis. IL-6, interleukin-6. TNF-α, tumor necrosis factor alpha. IL-1β, interleukin 1 beta. CXCL 12, chemokine ligand 12. hsCRP, high sensitivity C-reactive protein. GDF15, growth differentiation factor 15. HbA1c, glycated hemoglobin. CBC, complete blood count.

Kaplan-Meier curves comparing those who developed progressive CRS to those who did not show decreased survival in the progressive CRS group (*p* <0.001), **Figure 2**. Survival by subgroup of progressive CRS was also evaluated using Kaplan-Meier curves. Those with progressive CRS who developed HF followed by ESKD and those with progressive CRS who developed ESKD followed by HF had decreased survival when compared to those without progressive CRS (*p* <0.0001), **Figure 3A and 3B**. When the two subgroups were compared to each other there was no statistically significant difference seen in survival (*p* =0.25), **Figure 3C**.

**Figure 2.**
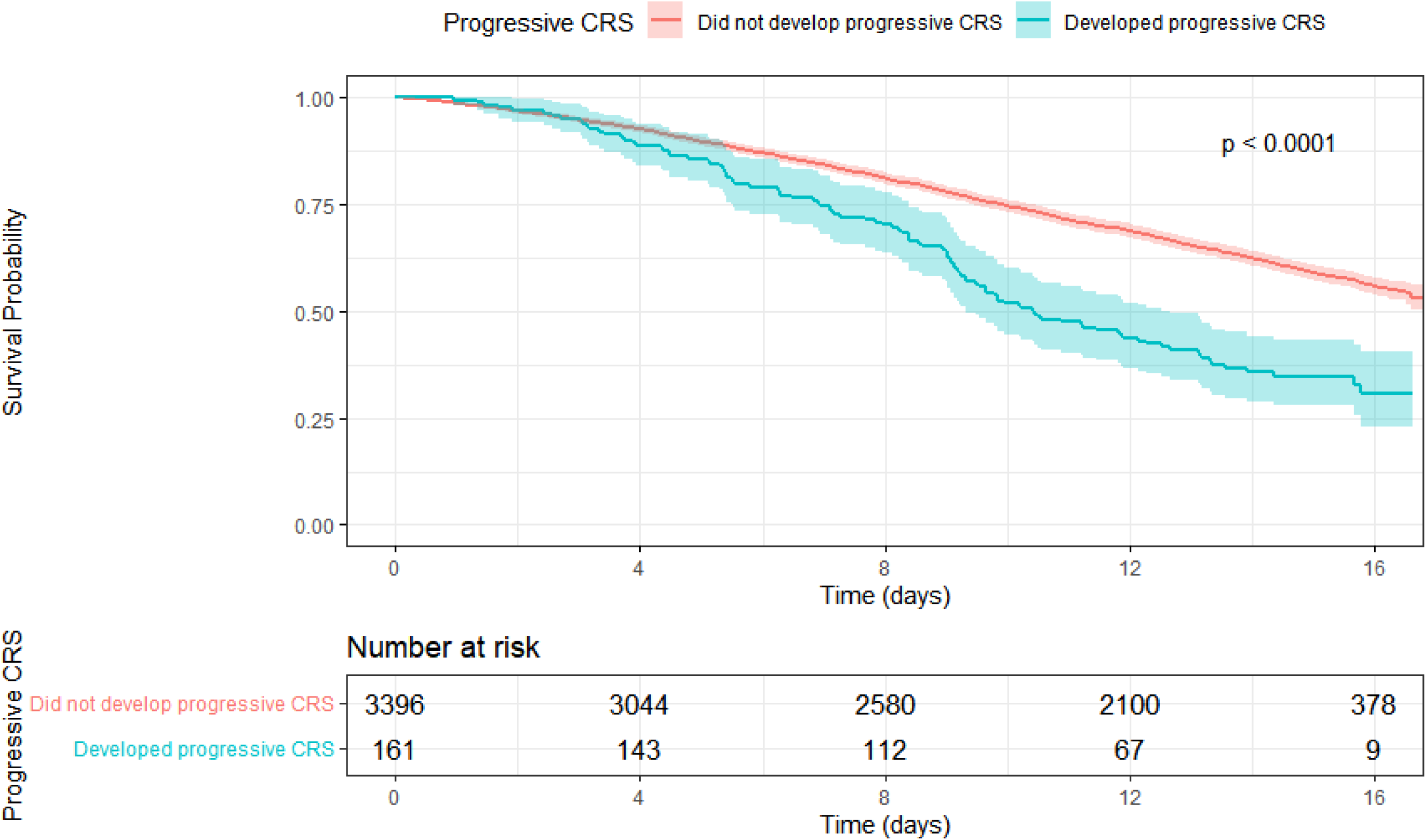
Kaplan-Meier Curve estimates of survival in those who developed progressive CRS vs those without development of progressive CRS.

**Figure 3.**
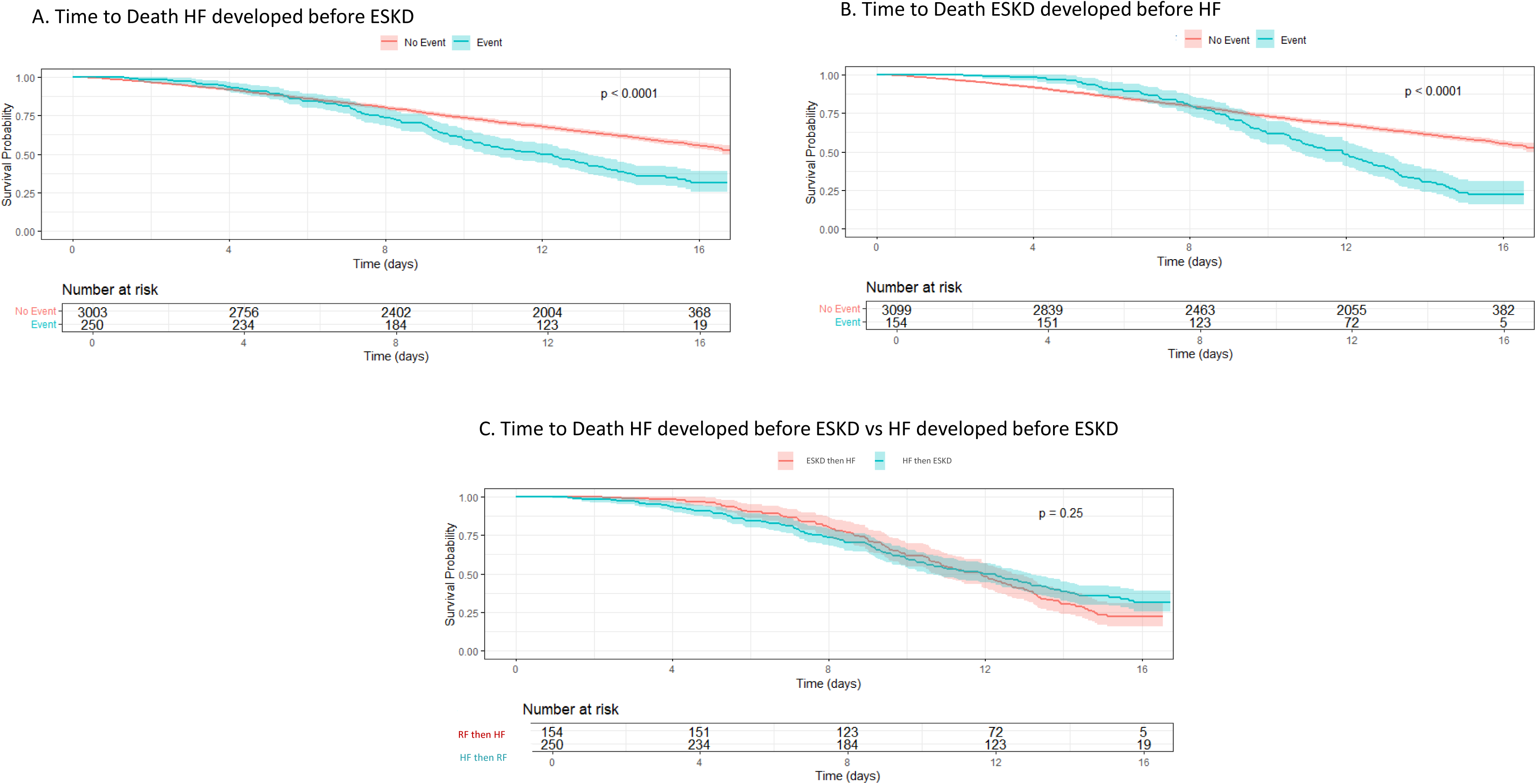
Kaplan-Meier Curve estimates of survival by progressive CRS subtype. Panel A shows survival comparison between those with progressive CRS where HF developed before ESKD compared to those who did not develop progressive CRS. Panel B shows survival comparison between those with progressive CRS where ESKD developed before HF compared to those who did not develop progressive CRS. Panel C shows survival comparison between those with progressive CRS who developed ESKD before HF compared to those with progressive CRS who developed HF before ESKD.

Associations of traditional risk factors with progressive CRS were also evaluated in cases where renal failure preceded heart failure, **Supplementary Table 1,** and when heart failure preceded renal failure, **Supplementary Table 2.** In multivariable-adjusted models, not graduating from high school, lower eGFR, and higher ACR were shared risk factors for both CRS subtypes. Current smoking was associated with progressive CRS among participants who developed renal failure before heart failure, whereas a history of clinical CVD, larger waist circumference, higher systolic blood pressure, and diabetes were associated with progressive CRS among those who developed heart failure before renal failure. The novel risk factors were also evaluated specifically in those who developed renal failure before heart failure, **Supplementary Table 3**, and those who developed heart failure before renal failure, **Supplementary Table 4**. Alkaline phosphatase, TNF-α, GDF-15, and hemoglobin were shared risk factors for both CRS subtypes. Higher PTH was associated with progressive CRS in the multivariable adjusted models for those who developed renal failure before heart failure. Whereas higher phosphate, calcium, FGF23, aldosterone, BNP, galactin 3, CTGF, IL-6, fibrinogen, CXCL, and hemoglobin A1C were associated with progressive CRS in those who developed heart failure followed by renal failure.

## Discussion

In this large, prospective study of adults with CKD, 4.5% developed a more progressive cardiorenal syndrome, defined as the development of both ESKD (requiring dialysis or renal transplant) and HF within 12 months. The incidence was higher among men and Hispanic individuals compared with their respective comparison groups. Individuals who developed progressive CRS have decreased survival compared to those who did not. However, there was no significant difference in survival between individuals with either HF proceeding ESKD or ESKD proceeding HF. This study finding also suggested that both traditional and non-traditional risk factors were associated with progressive CRS. Among traditional risk factors, less than high school education, history of CVD, lower eGFR, and higher albuminuria were strongly associated with development of progressive CRS. Additionally, one standard deviation higher of BNP, TNF-α, GDF-15, alkaline phosphatase, and hemoglobin A1C was associated with increased risk of progressive CRS while one standard deviation higher of hemoglobin was associated with decreased risk, independent of traditional risk factors.

These findings have important clinical and public health implications because CRS is highly prevalent and associated with significant high mortality among patients with CKD (1–5, 9). Understanding the potential etiology of CRS progression may guide the development of new approaches and more effective treatments to prevent CRS progression and its associated complications. The current study suggested that both traditional and non-traditional risk factors may play a role in CRS progression, targeting specific risk factors may represent a potential promising strategy.

The findings of this study suggest that several potentially modifiable traditional risk factors—such as current smoking, higher systolic blood pressure, higher triglyceride levels, lower eGFR, and higher ACR—may be associated with progressive CRS. Smoking has been linked to HF and an increased risk of CKD in previous studies (16,17). Both high blood pressure and elevated triglycerides are components of the Cardiovascular-Kidney-Metabolomic syndrome (18), and their association with progressive CRS requires further confirmation. In multivariable-adjusted models, the associations of smoking, blood pressure, and triglycerides with progressive CRS were attenuated. Only having less than a high school education, a history of clinical CVD, lower eGFR, and higher log-transformed ACR remained strongly associated with progressive CRS. A one standard deviation increases in eGFR and ACR was associated with a 36% lower risk or a 2.44-fold higher risk of progressive CRS, respectively, suggesting that preserving kidney function and reducing albuminuria may help prevent progressive CRS. These risk factors have been reported in a previous study of this cohort evaluating risk factors for incident heart failure, which found that having less than a high school education, a history of CVD, lower eGFR, and higher urine albumin were all independently associated with incident heart failure (15). Lower GFR has also been shown to correlate with an increased risk of CRS type 1 in other cohorts (16,17). Additionally, lower eGFR and higher albuminuria have been associated with higher all-cause and cardiovascular mortality (18). Taken together, patient education, preservation of eGFR, and reduction of albuminuria may serve as key strategies to prevent progressive CRS and warrant further evaluation.

We found that alkaline phosphatase, BNP, TNF-α, GDF-15, hemoglobin A1C, and hemoglobin were associated with progressive CRS. These findings suggest that in addition to some traditional CVD risk factors, bone mineral disease, fluid status, diabetes control, anemia, and inflammation are important risk factors for the development of progressive CRS. In the CRIC cohort, anemia, hemoglobin A1C, and TNF-α were also found to be associated with incident heart failure (19), while inflammatory marker CXCL12, NT-proBNP and kidney injury marker urinary NGAL were associated with CKD progression (20). Studies of other cohorts have also shown that elevated NT-proBNP is associated with rapid decline of kidney function (21), increased rates of ESKD (22), and AKI in those with HF (23,24). One study evaluating risk factors for CRS in elderly patients admitted with acute HF found that lower hemoglobin and hematocrit levels were predictive of developing CRS type 1 during admission (25). These findings suggest consistent associations of anemia, diabetes control, inflammation, and volume control with CRS and its progression. In our analysis GDF-15 was found to be associated with CRS. GDF-15 is a cytokine and growth factor that is secreted in response to cellular stress. GDF-15 has been shown to be predictive of worsening renal function in both AKI and CKD (26). Similarly, GDF-15 is prospectively associated with poor outcomes in patients with HF (27). Additionally, Alkaline phosphatase was independently associated with risk of CRS in our analysis. Alkaline phosphatase has previously been shown to be associated with mortality and worsening renal function in patients with kidney disease (28–30). Alkaline phosphatase is also associated with increased risk of cardiovascular disease (31) and mortality in those with existing cardiovascular disease (32). Among patients without cirrhosis, such as participants in the CRIC study, elevated alkaline phosphatase levels may reflect underlying bone disease, as approximately 40–50% of total alkaline phosphatase is derived from bone-specific isoenzymes. Our study suggests that control of these shared risk factors may prevent both CKD and heart failure. Additionally, prevention and treatment of mineral and bone disorder may reduce the progression of CRS.

Our sensitivity analysis showed that not graduating from high school, lower GFR, higher ACR, mineral bone disorder, higher TNF-α, higher GDF-15, and lower hemoglobin were associated with progressive CRS regardless of whether HF or ESKD developed first. These may be targetable shared risk factors among subtypes of CRS. Current smoking and higher PTH levels were associated with progressive CRS when ESKD preceded HF. In those who developed HF prior to ESKD, CVD, systolic BP, central obesity, diabetes, phosphate, calcium, FGF23, aldosterone, volume status, galectin 3, IL-6, fibrinogen, CXCL, and CTGF were more strongly associated with progressive CRS.

Our analysis reiterates that certain traditional risk factors are associated with heart failure and ESKD. We also add to the growing evidence that in addition to traditional risk factors other factors such as anemia, fluid status, inflammation, and bone mineral disease may be contributing to the progression of cardiac and renal disease in patients with CKD. These risk factors are present no matter which event, heart failure or renal failure, occurs first, which paves the way for targeted interventions to those at risk. Many of these traditional and novel risk factors may be modifiable, this represents a potential area of intervention for CKD patients to ultimately improve outcomes.

Our study has several strengths. We evaluated a diverse, large, multi-center cohort of CKD patients. We were able to include many covariables and biomarkers in our analysis. To our knowledge we are the first to describe characteristics, outcomes, and risk factors for individuals with CKD who develop both HF and ESKD within 12 months of each other. Some limitations are present as well. The event rate is relatively low which may limit statistical power. This study is observational which limits making inferences regarding causation.

In conclusion, our study showed that almost 5% of patients with underlying chronic kidney disease develop both HF and ESKD within 12 months. These individuals have increased mortality compared to those who do not develop progressive CRS. Both traditional and novel risk factors are associated with progressive CRS. Patient education, preservation of eGFR, and reduction of albuminuria may serve as key strategies to prevent progressive CRS. Additionally, mineral bone disorder, volume status, inflammation, anemia, and glycemic control may be modifiable risk factors for progressive CRS. Further studies are warranted to investigate whether controlling these risk factors may prevent CRS progression.

## Data Availability

CRIC data can be made available by reasonable request

## Disclaimer

A portion of the data reported here have been supplied by the United States Renal Data System (USRDS). The interpretation and reporting of these data are the responsibility of the author(s) and in no way should be seen as an official policy or interpretation of the U.S. government.

